# Relationship between COVID-19 death toll doubling time and national BCG vaccination policy

**DOI:** 10.1101/2020.04.06.20055251

**Authors:** Yutaka Akiyama, Takashi Ishida

## Abstract

In this manuscript, we showed a statistically significant difference of the doubling times of the death toll between the group of countries with national universal Bacillus Calmette-Guerin (BCG) vaccination and the group without it for recent years. Based on a statistical test, the distributions of the doubling time of these two groups were significantly different (p=0.007). Miller et al. reported the relationship between BCG vaccination and mortality for COVID-19 based on deaths per million inhabitants. However, they did not take into account the differences in COVID-19 detection rates among the countries and the epidemic stages of the countries. Therefore we used a doubling time of the death toll as a more stable indicator instead. We also investigated the dependency of the BCG strains. Among the 42 BCG-vaccinated countries, the median doubling time of the eight countries using “Tokyo 172-1” strain at least partially (Japan, Iraq, Malaysia, South Korea, Philippines, Saudi Arabia, Pakistan, and Bangladesh) was 7.2 days, and that of the other 34 vaccinated countries was 5.5 days. Their distributions were also significantly different (p=0.026).

---

The non-specific effects of Bacillus Calmette-Guerin (BCG) vaccination have been enthusiastically discussed.^1^ Miller et al. reported the relationship between BCG vaccination and mortality for COVID-19 based on deaths per million inhabitants.^2^ However they did not take into account the differences in COVID-19 detection rates among the countries and the epidemic stages of the countries. Therefore we used a doubling time (DT) of the death toll as a more stable indicator.

First, we investigated the DT of COVID-19 deaths in 57 countries (source: Our World in Data,^3^ as of 20 April 2020). We defined the baseline date of each country as the date when summed fatalities of at least ten people was first observed in the country. We covered all countries that have at least 10-day observation period and had a population^4^ of more than 1 million. To exclude the effect of social interventions, the death toll data of the country was truncated by 30 days after the baseline date (for 20 countries among 57).

Next, we surveyed the national BCG vaccination policies of each country (sources: BCG Atlas,^5^ WHO-UNICEF report,^6^ and other 20 papers^7-26^). The 57 countries were divided into two groups based on whether the majority of the population between the ages of 0 and 39 was vaccinated (“BCG”) or not (“non-BCG”). BCG vaccination records, especially cover rates, are reliable only for the last few decades for several countries. The BCG policy data collected as well as calculated DTs for 57 countries are shown in Table 1.

**Table 1.**
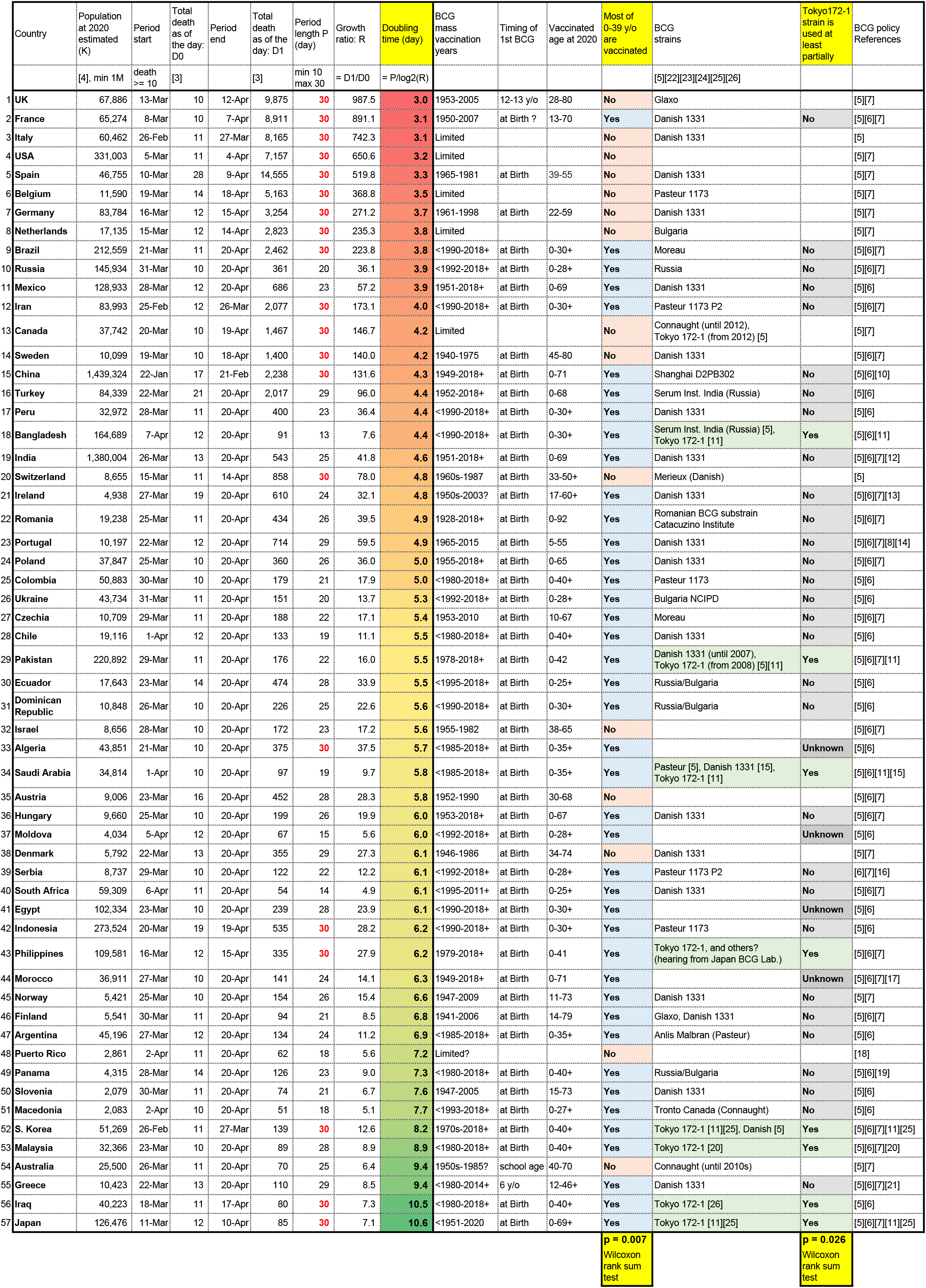
The COVID-19 death toll doubling time and the national BCG vaccination policy of 57 countries

Figure 1 shows the distributions of DTs. Most of DTs for 42 “BCG” countries are longer than 15 “non-BCG” countries (the medians of the DTs are 5.6 days and 4.2 days, respectively.) The variance of DTs of “BCG” countries was almost same as that of “non-BCG” (*σ* = 1.7 and *σ* = 1.8, respectively.) Based on a Wilcoxson rank-sum test, the distributions of these two groups were significantly different at the significance level of 0.05 *(p* = 0.007).

**Figure 1.**
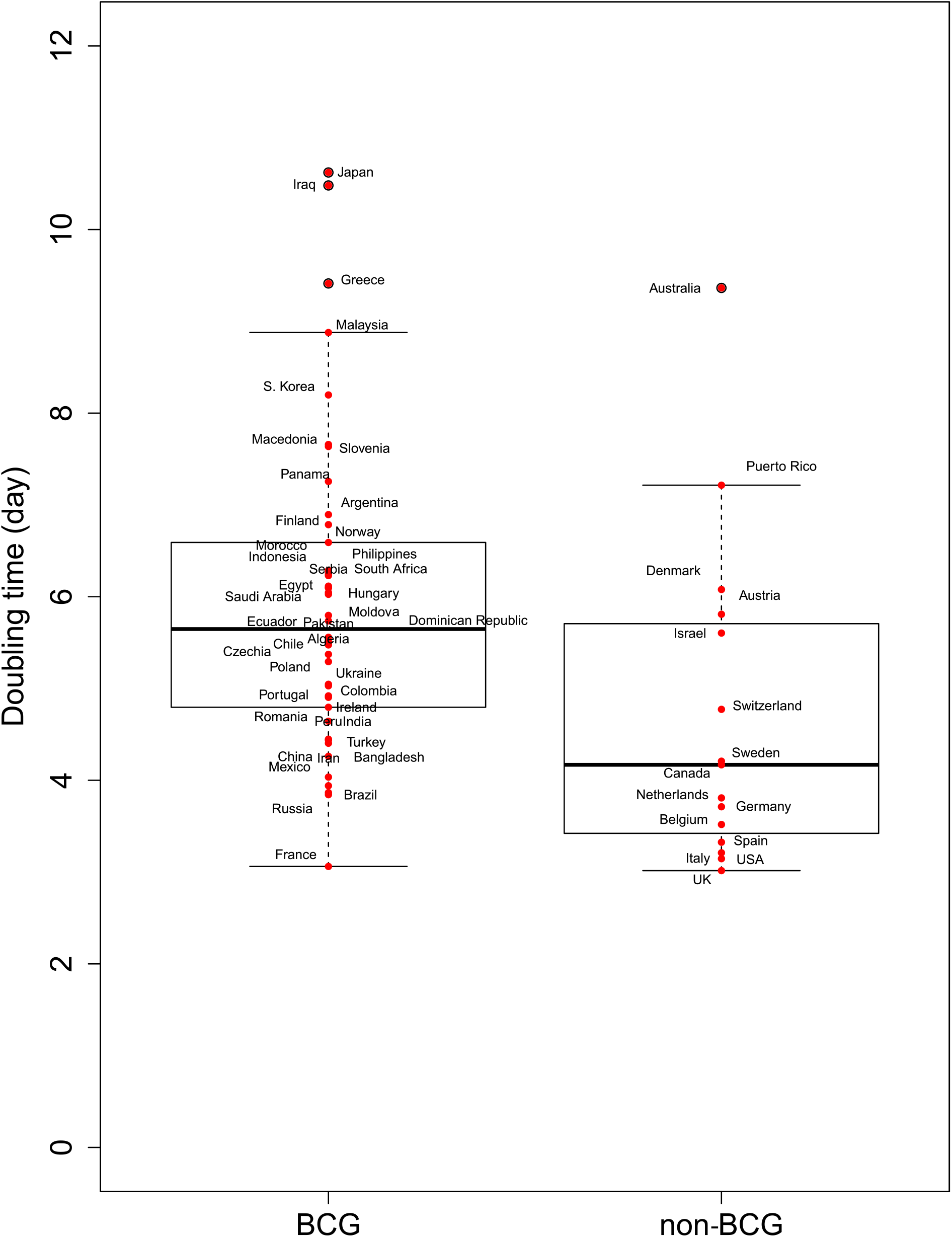
The death toll doubling time (DT) of BCG vaccinated (“BCG”) and non-vaccinated (“non-BCG”) countries

We also investigated the dependency of the BCG strains (Figure 2). Among the 42 “BCG” countries, the median DT of the eight countries using “Tokyo 172-1” strain at least partially (Japan, Iraq, Malaysia, South Korea, Philippines, Saudi Arabia, Pakistan, and Bangladesh) was 7.2 days, and that of the other 34 vaccinated countries was 5.5 days. The variance of DTs of “Tokyo 172-1” countries was large (*σ* = 2.4) while DTs of “other BCG strains” are relatively concentrated (*σ* = 1.3). Their distributions were also significantly different at the significance level of 0.05 (*p* = 0.026).

**Figure 2.**
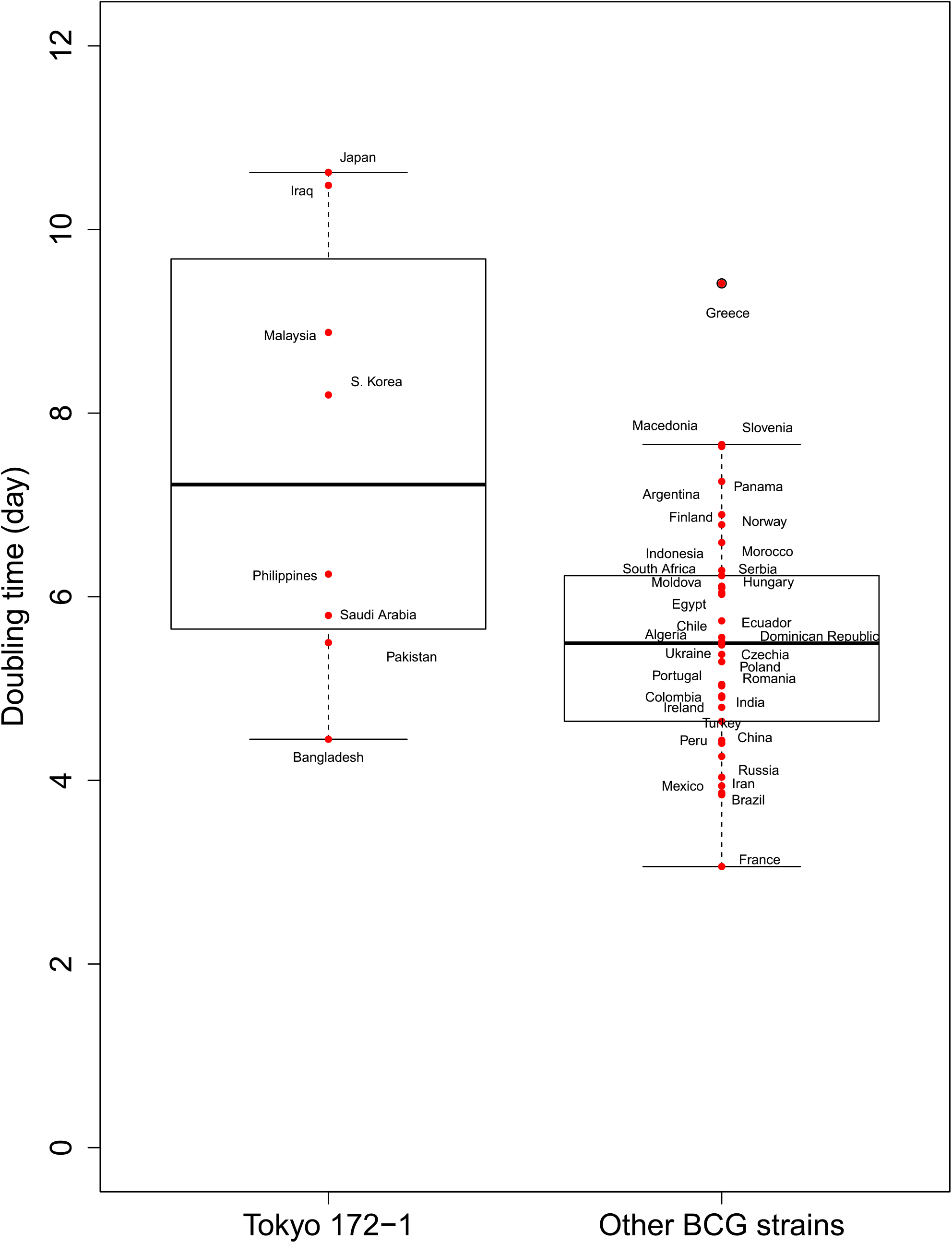
The death toll doubling time (DT) of Tokyo 172-1 (“Tokyo 172-1”) and other BCG strains (“Other BCG strains”) countries

We showed a statistically significant difference of the doubling times of the death toll between “BCG” and “non-BCG” countries. However, the correlation might be spurious and does not directly imply causality. In addition, we should carefully take the suggested difference between BCG strains, because the number of samples are not enough and some countries are using mixed strains.

The raw data version (an excel file) of Table 1 is available at http://www.bi.cs.titech.ac.jp/COVID-19/Death_vs_BCGpolicy.html.

## Authors’ contributions

Y.A. designed the study, and performed literature search, data table compilation, and basic data analysis. T.I. performed the statistical tests, and made figures. Y.A. and T.I. wrote the manuscript.

## Ethics committee approval

N/A

